# Obesity Modifies Clinical Outcomes of Right Ventricular Dysfunction

**DOI:** 10.1101/2023.01.18.23284734

**Authors:** Janet I. Ma, Emily Zern, Nona Jiang, Dongyu Wang, Paula Rambarat, Eugene Pomerantsev, Michael H. Picard, Jennifer E. Ho

## Abstract

**Introduction:** Right ventricular (RV) dysfunction is associated with increased mortality across a spectrum of cardiovascular diseases. The role of obesity in RV dysfunction and adverse outcomes is unclear.

**Methods:** We examined patients undergoing right heart catheterization between 2005-2016 in a hospital-based cohort. Linear regression was used to examine the association of obesity with hemodynamic indices of RV dysfunction [pulmonary artery pulsatility index (PAPi), right atrial pressure: pulmonary capillary wedge pressure ratio (RAP:PCWP), RV stroke work index (RVSWI)]. Cox models were used to examine the association of RV function measures with clinical outcomes.

**Results:** Among 8285 patients (mean age 63 years, 40% women), higher BMI was associated with worse indices of RV dysfunction, including lower PAPi (β -0.26, SE 0.01, p <0.001), higher RA:PCWP ratio (β 0.25, SE 0.01, p-value <0.001), and lower RVSWI (β -0.05, SE 0.01, p-value <0.001). Over 7.3 years of follow-up, we observed 3006 mortality and 2004 heart failure (HF) hospitalization events. RV dysfunction was associated with greater risk of mortality (eg PAPi: HR 1.11 per 1-SD increase, 95% CI 1.04-1.18), with similar associations with risk of HF hospitalization. BMI modified the effect of RV dysfunction on outcomes (P-interaction <=0.005 for both), such that the effect of RV dysfunction was more pronounced at higher BMI.

**Conclusions:** Patients with obesity had worse hemodynamic measured indices of RV function across a broad hospital-based sample. While RV dysfunction was associated with worse clinical outcomes including mortality and HF hospitalization, this association was especially pronounced among individuals with higher BMI.

## INTRODUCTION

The right ventricle has been historically overshadowed by its counterpart, the left ventricle, in its clinical significance across a broad spectrum of cardiovascular disease and particularly in heart failure. This is in part due the difficulty of understanding the role that right ventricular (RV) dysfunction plays in cardiovascular disease, both anatomically and physiologically [1]. Non-invasive measurements using echocardiography and cardiac magnetic resonance imaging to characterize function are limited by the RV’s unique anatomy, which is highly sensitive to different pressure and volume conditions.

From a physiology standpoint, RV dysfunction is intimately related to the pulmonary circulation and is variably affected by a wide range of cardiovascular and pulmonary disease phenotypes. This leads to a heterogenous population with RV dysfunction, which results in challenges in studying the subsequent impact that RV dysfunction may have on adverse outcomes [1,2].

Obesity is a growing problem in the United States with well-recognized contributions to incident cardiovascular disease and heart failure [3, 4]. While obesity has been associated with adverse RV remodeling using echocardiography and cardiac magnetic resonance measurements [5, 6], the association of obesity with invasive hemodynamic indices of RV function that may more accurately reflect hemodynamic consequences of RV function remains unclear. We leveraged a large hospital-based sample of individuals spanning a broad spectrum of cardiopulmonary disease who had undergone clinically indicated right heart catheterization. This provided a unique setting to examine the association of obesity with hemodynamic indices of RV function, and the association of RV function with subsequent clinical outcomes. We hypothesized that obesity is associated with RV dysfunction, and that obesity modifies the association of RV dysfunction with adverse clinical outcomes.

## METHODS

### Study Sample

We examined consecutive ambulatory and hospitalized patients undergoing right heart catheterization (RHC) between 2005 and 2016 at Massachusetts General Hospital. For patients who had multiple RHC procedures during this time period, only the initial RHC was included for analysis, resulting in a total of 10,306 cases. The following clinical exclusion criteria were applied: acute myocardial infarction occurring on the same day as catheterization, cardiac arrest or shock within 24 hours, presence of mechanical ventilation, presence of intra-aortic balloon pump, history of heart or lung transplant, complicated adult congenital heart disease, history of valvular replacement, or those on dialysis (n=887 excluded). Cases were also excluded if there were missing key clinical covariates (n=484), patient identifier variables (n=398), or hemodynamic parameters (n=252), leading to a final study sample of 8285 for analysis. This study was approved by the Mass General Brigham Institutional Review Board.

### Clinical and RV hemodynamic variables

Clinical characteristics were ascertained from the medical records at the time of RHC, including age, sex, body mass index (BMI), smoking status, and presence of comorbidities (diabetes mellitus, hypertension, history of myocardial infarction, history of heart failure, prior lung disease, and chronic kidney disease). Obstructive sleep apnea was identified by the electronic medical record utilizing appropriate *International Classification of Diseases Ninth Revision* (*ICD*-*9*) or *Tenth Revision* (*ICD*-*10*) codes.

Hemodynamic measures were recorded at the time of RHC, including resting blood pressure, heart rate, mean right atrial (RA) pressure, pulmonary artery (PA) systolic and diastolic pressure, and mean pulmonary capillary wedge pressure (PCWP). Nonphysiologic parameters were set to missing. The PA pulsatility index (PAPi) was calculated as 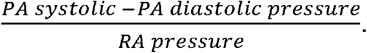. In the cases that RA pressure was recorded as zero, the value was set to one for the purposes of PAPi calculation, otherwise the RA pressure was set to missing for other non-physiologic values. Cardiac output and index were derived via thermodilution methods utilizing the Mosteller formula for body surface area^22^. RVSWI was derived as *0*.*0136 x Stroke volume index x (Mean PA pressure - mean RA pressure)* utilizing the thermodilution cardiac index measurement.

### Clinical Outcomes

All-cause mortality was ascertained using the National Social Security Death Master Index and hospital records, abstracted on 06/10/2020. Due to confidentiality purposes, the precise dates of deaths that occurred between 06/10/2017 and the date of abstraction are protected nationally. To conduct time-to-event analyses, these death dates were imputed at the midpoint of the blanking period (12/10/2018). Occurrence of a major adverse cardiac event (MACE) was defined as a composite of heart failure (HF) admission, cerebrovascular accident (CVA), transient ischemic attack (TIA) or acute myocardial infarction (MI) with a corresponding ICD-9 or ICD-10 code as the primary discharge diagnosis. A heart failure hospitalization was defined by an ICD-9 or ICD-10 code for heart failure as the primary discharge diagnosis or a current procedural terminology (CPT) code for heart transplantation (OHT) or durable ventricular assist device (VAD). The follow-up period for each participant was defined as time from RHC to death date or date of final encounter in the electronic health record. Patients were censored based on time of last encounter.

### Statistical Analysis

Baseline characteristics were summarized across the total sample and according to obesity class (Normal = BMI <25, Overweight = BMI ≥25 and <30, Obesity class 1 = BMI ≥30 and <35, Obesity class 2+3 = BMI ≥35 kg/m^2^). We examined the cross-sectional association of BMI and obesity class with hemodynamic indices of RV dysfunction, including PAPi, RA:PCWP ratio, and RVSWI using multivariable linear regression. In order to limit leverage of outliers in the analyses, the distribution of PAPi was winsorized by setting the minimum PAPi as 0.3 and the maximum PAPi as 30. PAPi, RA:PCWP ratio, and RVSWI were natural log-transformed due to right-skewed distributions. Models were adjusted for the following clinical covariates: age, sex, hypertension, diabetes mellitus, obstructive sleep apnea (OSA), chronic lung disease, pulmonary hypertension, prevalent MI, and prevalent HF. Further, we used least squares means to estimate adjusted mean values for untransformed RV function indices across obesity classes after controlling for all covariates.

To investigate the effect of obesity and RV dysfunction and their potential interaction on future clinical outcomes, we first examined the association of RV function measures known to be associated with obesity from primary analyses with clinical outcomes, including all-cause mortality and HF hospitalizations. We used the Kaplan-Meier method to examine the association of RV function measures with outcomes and log rank tests to examine differences between groups. Multivariable Cox models were constructed, adjusting for age, sex, BMI, previous MI, previous HF, the presence of pulmonary hypertension (PH, defined as mean pulmonary arterial pressure ≥20), hypertension, diabetes, and chronic kidney disease. The proportional hazards assumption was tested, and minor violations were found using cumulative Martingale residuals. We therefore included time-varying predictors to account for model fitness, including log-transformed PAPi, age, BMI, previous HF, and hypertension treatment.

We then examined whether BMI modified the association of RV function with clinical outcomes. We used multiplicative interaction terms (PAPi*BMI and RA:PCWP ratio*BMI) entered into multivariable Cox models to examine this. Further, to illustrate effect modification, we plotted estimated adjusted hazard ratio of each RV function measure with clinical outcome (y-axis) by BMI (x-axis). A two-sided p value <0.05 was deemed as statistically significant. Analyses were conducted using SAS software, Version 9.4 (Cary, North Carolina, USA).

## RESULTS

We studied 8285 individuals with a mean age of 63 ± 13 years, 39% of whom were women. Comorbid conditions were common including 59% with hypertension, 23% with diabetes mellitus, 16% with chronic lung disease, 14% with obstructive sleep apnea, 19% with previous MI, and 32% with previous HF. Baseline characteristics stratified by obesity class are shown in **Table 1**. Mean BMI across the sample was 29.3 ± 6.8 kg/m^2^, and 28% of participants were classified as normal weight, 34% overweight, 21% obesity class 1, and 17% obesity class 2+3. Patients across increasing obesity classes were similar in age, with greater comorbid burden including hypertension, diabetes, prior HF, and obstructive sleep apnea. Of note, natriuretic peptide levels were lower across obesity classes despite higher right- and left-sided filling pressures (**Table 1**).

**Table 1.** Baseline characteristics by obesity class

|  | **Normal**<br>**(n=2307)** | **Overweight**<br>**(n=2793)** | **Obese Class 1**<br>**(n=1732)** | **Obese Class**<br>**2+3**<br>**(n=1453)** | **p-**<br>**value** |
| --- | --- | --- | --- | --- | --- |
| **Clinical characteristics** |  |  |  |  |  |
| Age, years | 62 ± 15 | 64 ± 12 | 64 ± 11 | 62 ± 12 | <0.001 |
| Female, n (%) | 1111 (48.2) | 877 (31.4) | 624 (36) | 651 (44.8) | <0.001 |
| Race, n (%) |  |  |  |  | <0.001 |
| White | 1900 (82.4) | 2377 (85.1) | 1518 (87.6) | 1234 (84.9) |  |
| Black | 88 (3.8) | 97 (3.5) | 58 (3.4) | 69 (4.8) |  |
| Hispanic | 75 (3.3) | 98 (3.5) | 46 (2.7) | 43 (3.0) |  |
| Asian | 86 (3.7) | 54 (1.9) | 18 (1) | 6 (0.4) |  |
| Unknown | 158 (6.9) | 167 (6) | 92 (5.3) | 101 (7) |  |
| BMI, kg/m<sup>2</sup> | 22.2 ± 2.1 | 27.5 ± 1.4 | 32.3 ± 1.4 | 40.5 ± 5.4 | <0.001 |
| Systolic BP, mmHg | 124.5 ± 25.7 | 124.8 ± 23.8 | 128.6 ± 24.3 | 130.2 ± 23.6 | <0.001 |
| Heart rate, bpm | 73.9 ± 16 | 71.6 ± 15.2 | 71.4 ± 14.5 | 73.8 ± 15.5 | <0.001 |
| Hypertension, n (%) | 1052 (45.6) | 1675 (60) | 1150 (66.4) | 996 (68.6) | <0.001 |
| Diabetes Mellitus, n (%) | 286 (12.4) | 596 (21.3) | 505 (29.2) | 536 (36.9) | <0.001 |
| Previous MI, n (%) | 361 (15.7) | 563 (20.2) | 373 (21.5) | 254 (17.5) | <0.001 |
| Previous HF, n (%) | 706 (30.6) | 860 (30.8) | 587 (33.9) | 503 (34.6) | 0.010 |
| Hypercholesterolemia, n (%) | 1078 (62.9) | 1774 (76.7) | 1151 (80.7) | 974 (84.4) | <0.001 |
| Chronic lung disease, n (%) | 347 (15) | 416 (14.9) | 319 (18.4) | 267 (18.4) | 0.001 |
| OSA, n (%) | 101 (4.4) | 239 (8.6) | 302 (17.4) | 512 (35.2) | <0.001 |
| Chronic kidney disease, n (%) | 54 (2.3) | 85 (3) | 64 (3.7) | 38 (2.6) | 0.070 |
| Current smoker, n (%) | 138 (10.6) | 131 (8.4) | 110 (11.2) | 81 (9.4) | 0.07 |
| BNP, pg/ml | 2928 (1031, 7346) | 2089 (633, 4997) | 1273 (402, 3177) | 991 (297, 2576) | <0.001 |
| **Hemodynamics** |  |  |  |  |  |
| RA mean pressure, mmHg | 5 (3, 8) | 6 (4, 9) | 8 (5, 11) | 10 (7, 14) | <0.001 |
| PA systolic pressure, mmHg | 34 (26, 46) | 35 (27, 47) | 36 (30, 49) | 41 (33, 53) | <0.001 |
| PA diastolic pressure, mmHg | 13 (8, 19) | 13 (9, 20) | 15 (10, 21.5) | 18 (12, 25) | <0.001 |
| PCWP, mmHg | 12 (8, 18) | 13 (9, 19) | 15 (10.5, 20) | 17 (12, 24) | <0.001 |
| Td cardiac output, L/min | 4.5 (3.6, 5.5) | 5 (4.2, 6) | 5.2 (4.4, 6.3) | 5.8 (4.8, 7) | <0.001 |
| PAPi | 4.4 (2.8, 7.8) | 3.6 (2.4, 5.8) | 3 (2, 4.5) | 2.5 (1.7, 3.6) | <0.001 |
| RA:PCWP ratio | 0.2 (0.1, 0.3) | 0.3 (0.2, 0.4) | 0.3 (0.2, 0.4) | 0.4 (0.3, 0.4) | <0.001 |
| RVSWI, gm-m/m2/beat | 7.9 (5.8, 11.1) | 8 (5.9, 11) | 8.1 (6.1, 11.3) | 8.4 (6.1, 11.7) | 0.010 |
| Pulmonary hypertension, n(%) | 1261 (54.7) | 1630 (58.4) | 1162 (67.1) | 1155 (79.5) | <0.001 |

Table displays mean ± standard deviation. \* denotes variables displayed as median [interquartile range].
Abbreviations: PAPi, pulmonary artery pulsatility index; MI, myocardial infarction; HF, heart failure; OSA, obstructive sleep apnea; BMI, body mass index; NT-proBNP, N-terminal pro-B type natriuretic peptide; PA, pulmonary artery; RVSWI, right ventricular stroke work index (calculated using thermodilution cardiac output), Td, thermodilution method. RVSWI was calculated by thermodilution cardiac output measurement.

### BMI and Obesity are Associated with Worse RV function

Higher BMI was associated with worse indices of RV function across all three measures. Specifically, PAPi was lower across obesity classes (4.4 [2.8, 7.8] in normal weight vs 2.5 [1.7, 3.6] in class 2+3 obesity), and RA:PCWP ratio was higher across obesity classes (0.2 [0.1, 0.3] in normal weight vs 0.4 [0.3, 0.4] in class 2+3 obesity). Lastly, RVSWI was lower among those with higher BMI, though there were minimal differences across obesity classes. When applying the clinical cutpoint of PAPi<2 to define RV dysfunction based on previous studies, 12% individuals with normal weight, 16% overweight, 22% class I, and 32% class 2+3 obesity had RV dysfunction. [20] (**Table 2**).

**Table 2.** Associations of BMI and obesity classes with indices of RV function

| Predictors | $\beta$ estimate | Standard error | p value | p-trend |
| --- | --- | --- | --- | --- |
| ***PAPi*** |  |  |  |  |
| BMI | -0.23 | 0.01 | <0.001 | N/A |
| Obesity Class |  |  |  |  |
| Normal | REF |  |  |  |
| Overweight | -0.25 | 0.03 | <0.001 | <0.001 |
| Class 1 | -0.48 | 0.03 | <0.001 |  |
| Class 2+3 | -0.65 | 0.03 | <0.001 |  |
| ***RA:PCWP ratio*** |  |  |  |  |
| BMI | 0.25 | 0.01 | <0.001 | N/A |
| Obesity Class |  |  |  |  |
| Normal | REF |  |  |  |
| Overweight | 0.28 | 0.03 | <0.001 | <0.001 |
| Class 1 | 0.51 | 0.03 | <0.001 |  |
| Class 2+3 | 0.69 | 0.03 | <0.001 |  |
| ***RVSWI*** |  |  |  |  |
| BMI | -0.05 | 0.01 | <0.001 | N/A |
| Obesity Class |  |  |  |  |
| Normal | REF |  |  |  |
| Overweight | -0.02 | 0.03 | 0.47 | 0.88 |
| Class 1 | -0.08 | 0.03 | 0.01 |  |
| Class 2+3 | -0.15 | 0.03 | <0.001 |  |

$\beta$ estimates represent changes in natural log-transformed outcomes per 1-SD increase in BMI. p-trend demonstrates the linear trend across BMI groups. Multivariable models were adjusted for age, sex, hypertension, diabetes mellitus, OSA, chronic lung disease, prevalent MI, prevalent HF, and PH.

In multivariable-adjusted analyses, we found that higher BMI was associated with worse indices of RV function, including lower PAPi, higher RA:PCWP ratio, and lower RVSWI (**Table 2**). Specifically, we found that a 1-SD higher BMI was associated with a 0.26 log-unit lower PAPi (β -0.3, SE 0.01, p <0.001). In adjusted analyses using least squares means, the mean PAPi among normal weight individuals was 4.5 vs 2.8 among those with class 2+3 obesity (**Figure 2**). Similarly, a 1-SD higher BMI was associated with a 0.25 log-unit higher RA:PCWP ratio (β 0.25, SE 0.01, p-value <0.001), and mean RA:PCWP ratio among normal weight individuals was 0.21 vs 0.32 among those with class 2+3 obesity (**Figure 2**). Lastly, a 1-SD higher BMI was associated with a 0.05 log-unit lower RVSWI (β -0.05, SE 0.01, p-value <0.001). Mean RVSWI in normal weight individuals was 8.26 vs 7.66 in those with class 2+3 obesity.

**Figure 1.**
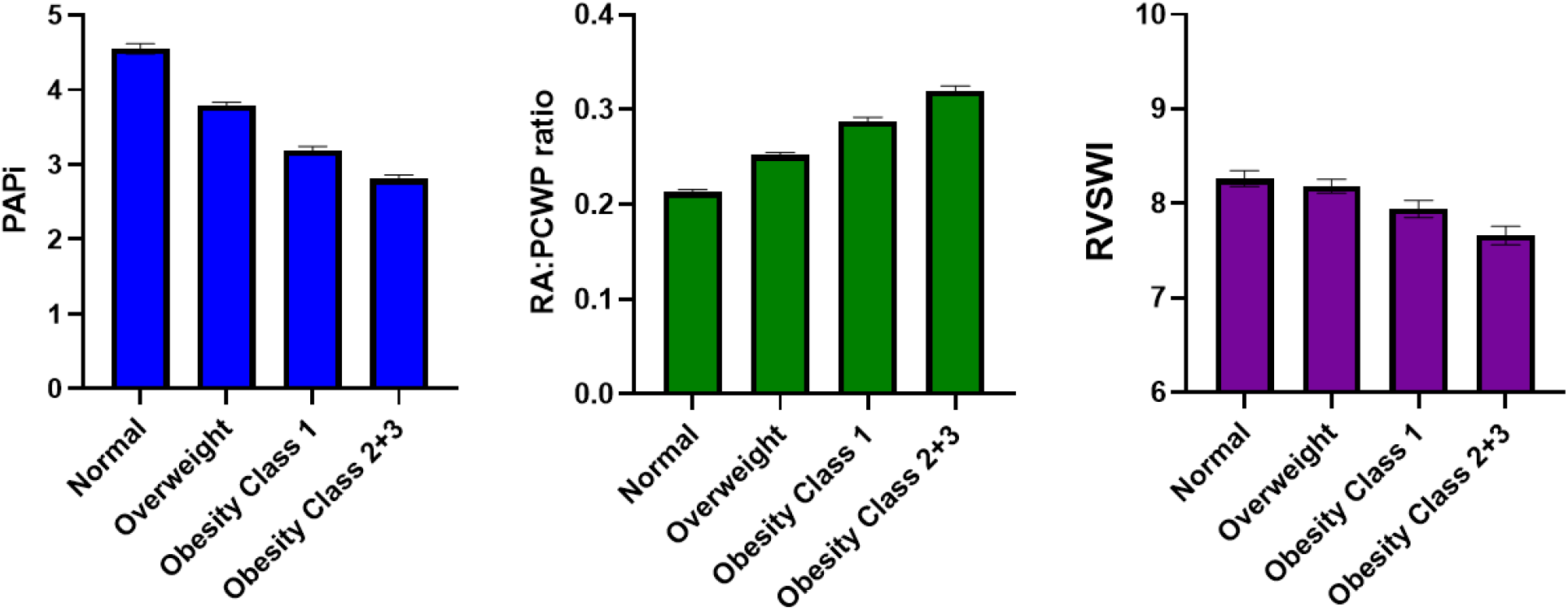
Hemodynamic indices of RV function across obesity classes Least square means represent adjusted mean outcome for each obesity category listed in the table after controlling for all covariates. Multivariable models were adjusted for age, sex, hypertension, diabetes mellitus, OSA, chronic lung disease, prevalent Ml, and prevalent HF. BMI categories: Normal = BMI <25, Overweight = BMI >25 and <30, Obesity class 1 = BMI >30 and <35, Obesity class 2+3 = BMI >35.

**Figure 2.**
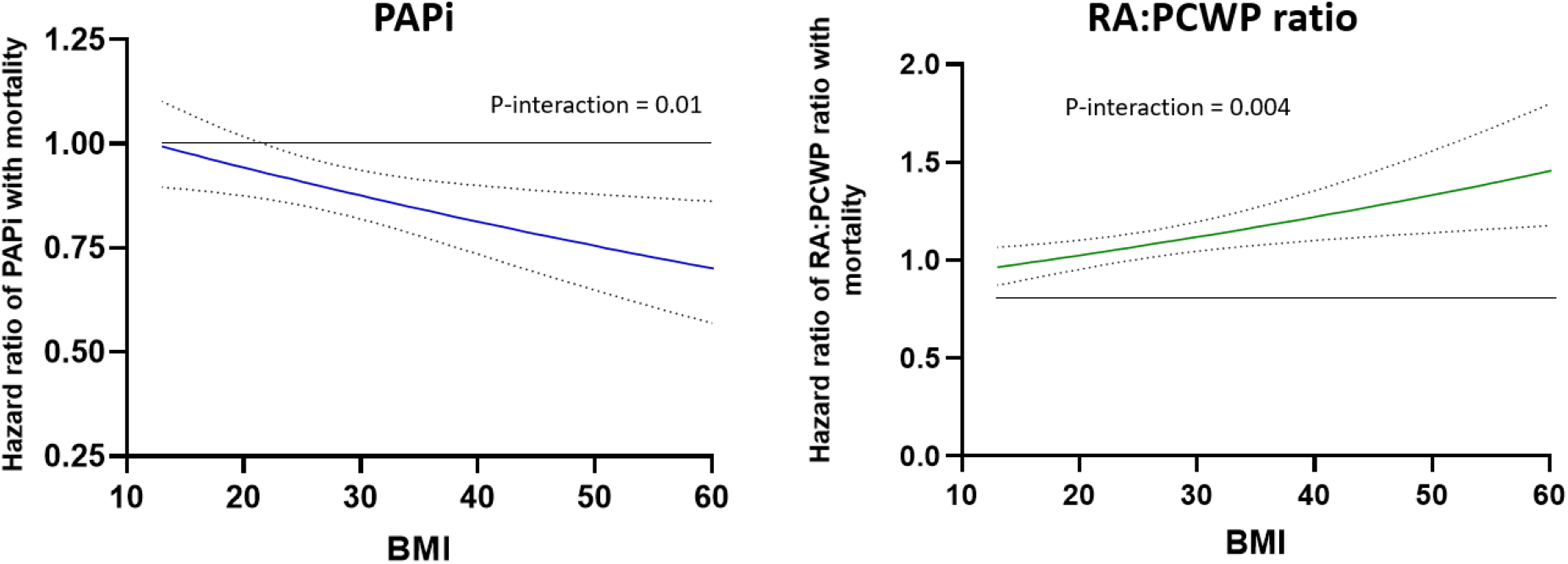
BMI modifies the effect of RV function on mortality HR represents effect on mortality per 1-SD change in log-transformed PAPi or RA:PCWP ratio. Multivariable model adjusted for age, sex, hypertension, diabetes, prior myocardial infarction, and prior heart failure. Abbreviations: BMI – body mass index, HR – hazard ratio, PAPi – pulmonary artery pulsatility index, RA:PCWP – right atrium: pulmonary capillary wedge pressure, SD – standard deviation

Given known associations of BMI and RV function with PH as well as HF, we examined whether PH or HF status may modify the effect of BMI on RV dysfunction in exploratory analyses. Whereas PH did not modify the effect of BMI or obesity on PAPi or RA:PCWP ratio (P for interaction >0.30 for both), we did find that the effect of BMI on RVSWI was more pronounced among those without vs with PH (P_int_ = 0.02 in multivariable analyses). In stratified analyses, a 1-SD higher BMI was associated with 0.08 lower RVSWI (beta -0.08, SE 0.02, p-value <0.001) among those without PH, compared with 0.04 lower RVSWI among those with PH (beta -0.08, SE 0.02, p-value <0.001). We did not find that HF status modified the effect of BMI on RV dysfunction.

### Obesity and RV Function Measures are Associated with Clinical Outcomes

Over 7.3 years of follow-up, we observed 3006 mortality and 2004 heart failure hospitalization events, with Kaplan-Meier curves by obesity class shown in **Supplemental Figure 1**.

We found that indices of RV function were associated with adverse outcomes in multivariable-adjusted analyses. Specifically, a 1-SD higher PAPi was associated with a 10% lower hazard of all-cause mortality, and a 1-SD higher RA:PCWP ratio was associated with an 8% higher hazard (PAPi: HR 0.90, 95% CI 0.84-0.96; RA:PCWP ratio: HR 1.08 per 1-SD increase, 95% CI 1.02-1.15), with similar associations with HF hospitalization (PAPi: HR 0.79, 95% CI 0.74-0.84; RA:PCWP ratio: HR 1.14 per 1-SD increase, 95% CI 1.07-1.22). Interestingly, while a 1-SD higher RVSWI was associated with a 12% lower hazard of HF hospitalization (HR 0.88, CI 0.83-0.94), it was also associated with a 6% higher hazard of all-cause mortality (HR 1.06, CI 1.02-1.11). (**Table 3)**

**Table 3.** Associations between adverse outcomes and RV function

| Predictors | Multivariable Adjusted |  |  |  |
| --- | --- | --- | --- | --- |
|  | HR | 95% CI | p value | p-int<sub>BMI</sub> |
| **All-Cause Mortality** |  |  |  |  |
| PAPi | 0.84 | 0.78-0.89 | <0.001 | 0.001 |
| RA:PCWP ratio | 1.11 | 1.04-1.18 | 0.001 | <0.001 |
| RVSWI | 1.26 | 1.21-1.32 | <0.001 | 0.50 |
| **HF hospitalization** |  |  |  |  |
| PAPi | 0.72 | 0.67-0.77 | <0.001 | 0.06 |
| RA:PCWP ratio | 1.17 | 1.10-1.25 | <0.001 | 0.17 |
| RVSWI | 1.13 | 1.06-1.20 | <0.001 | 0.03 |

Hazard ratios (HR) represent incremental risks per 1-SD increase in the predictors. P-int<sub>BMI</sub> shows the significance of multiplicative interaction between predictors and BMI. Multivariable models were adjusted for age, sex, hypertension, diabetes mellitus, OSA, chronic lung disease, prevalent MI, prevalent HF, and PH.

Moreover, we found that BMI modified the effect of RV function on both mortality (P-interaction <=0.005) and heart failure hospitalizations (P-interaction <=0.005) as demonstrated in **Figure 3**. Specifically, the association of lower PAPi with worse clinical outcomes was more pronounced at higher BMI. For example, at BMI of 20 kg/m^2^, a 1-SD lower PAPi was associated with a 6% higher hazard of mortality (HR 1.06, 95% CI 0.98-1.1), whereas at a BMI of 40 kg/m^2^, a 1-SD lower PAPi was associated with a 23% higher hazard of mortality (HR 1.23, 95% CI 1.1-1.4). Similarly, the association of higher RA:PCWP ratio with greater mortality was more pronounced at higher BMI. For example, at BMI of 20 kg/m^2^, a 1-SD higher RA:PCWP ratio was associated with a 3% higher hazard of mortality (HR 1.03, 95% CI 0.95-1.1), whereas at a BMI of 40 kg/m^2^, a 1-SD higher RA:PCWP ratio was associated with a 22% higher hazard of mortality (HR 1.22, 95% CI 1.1-1.35).

## DISCUSSION

We investigated the association of obesity and BMI with hemodynamic indices of RV function in a hospital-based sample representing a broad spectrum of cardiopulmonary disease referred for RHC. We found that obesity and higher BMI were consistently associated with hemodynamic measures of RV dysfunction. More importantly, measures of RV dysfunction were associated with an increased risk of mortality and heart failure hospitalization, and obesity significantly heightened the risk RV dysfunction conferred on adverse outcomes. Taken together, these findings underscore the importance of obesity as a risk enhancing factor for RV failure, with important implications on subsequent adverse outcomes. Our findings are of particular interest considering rapidly evolving therapies for obesity, including glucagon-like peptide-1 receptor agonists or dual glucose-dependent insulinotropic polypeptide and glucagon-like peptide-1 receptor agonists, with potential future implications on the prevention of cardiovascular disease that remain to be elucidated [7,8].

Prior studies have examined the association of obesity and non-invasive measures of RV structure and function using echocardiography and magnetic resonance imaging (MRI). For example, overweight and obese individuals had lower RV free wall strain and strain rate as well as greater RV free wall thickness and volume as measured by echocardiography compared with individuals with normal BMI in a modest sized case-control study [9]. These findings were corroborated in a larger retrospective study, which found higher BMI was independently associated with abnormal RV free wall longitudinal strain in over one thousand individuals [10]. Similar findings have been observed in the MESA-Right Ventricle study, a cardiac MRI-based study where overweight and higher obesity classes were associated with greater RV mass, larger RV volumes, and slightly lower RV ejection fraction [11]. However, accurate standard echocardiographic assessment is challenged by the complex geometry of the RV, and CMR has practical limitations to widespread use.

Our study adds to previous findings by demonstrating that higher BMI is associated with hemodynamic measures of RV dysfunction including lower PAPi, lower RVSWI, and higher RA:PCWP ratio independent of clinical cardiopulmonary disease. Indeed, when stratified by pre-existing heart failure or PH, we found that higher BMI was still independently associated with lower PAPi and higher RA:PCWP ratio. In this context, utilizing invasive hemodynamic indices of RV function is a strength of our study, allowing for more direct physiologic assessment and leveraging parameters with prognostic implications. Low PAPi has previously been shown to be associated with higher mortality in select samples including post-myocardial infarction, pulmonary arterial hypertension, and advanced heart failure populations [12-17]. We recently demonstrated that lower PAPi was associated with increased risk of adverse events including all-cause mortality, MACE, and HF hospitalizations across a much wider spectrum of cardiopulmonary disease [18]. Similarly, RA:PCWP ratio as an index for RV function has also been demonstrated as a marker for in-hospital mortality [19]. Our results of PAPi and RA:PCWP ratio are in keeping with these prior studies but now extend findings across a broader sample of individuals.

Interestingly, our findings related to RVSWI showed a slightly higher hazard of all-cause mortality on one hand, along with a lower hazard of HF hospitalization. Traditionally, RVSWI has been used to assess risk of RV failure in heart transplantation as well as need for mechanical right-sided support following implantation of left ventricular assist devices [20, 21]. However, other populations appear to have more heterogenous clinical outcomes associated with RVSWI. In a retrospective, multi-center registry study examining RV dysfunction in cardiogenic shock from either acute MI or HF, while individuals with greater shock had lower RVSWI consistent with worse RV dysfunction, RVSWI was significantly associated with mortality in the HF cohort but not in the acute MI cohort [22]. Another cross-sectional study found that a higher RVSWI was associated with worse renal function in individuals with HFpEF, which was attributed to the presence of fixed pre-capillary PH in a sub-group leading to increased RV afterload [23]. Given the diverse spectrum of individuals studied in our sample, our seemingly contradictory findings may be due to these same heterogeneous associations of RVSWI with outcomes across different disease states.

Further, we demonstrate that the association of RV dysfunction with adverse outcomes is particularly pronounced at higher BMI. This highlights the importance of obesity as an important effect modifier when considering clinical implications of RV dysfunction. Prior small studies have suggested that adverse RV remodeling may be reversible with weight loss. One study examining echocardiographic parameters of adverse cardiac remodeling in 30 individuals with a starting BMI of >30 suggested that right ventricular dysfunction along with LV diastolic function may be reversible with weight loss over a 3-month period [24]. Another study of 15 females with obesity but no cardiovascular disease found an association with right ventricular dysfunction in obesity and an impaired functional capacity, both which recovered after consistent weight loss [25]. Building on these studies, our results therefore emphasize the potential for greater clinical impact that RV dysfunction has on individuals with higher BMI than previously appreciated.

The mechanisms underlying the relationship between RV dysfunction and obesity remain unclear and are likely multifactorial. This includes a number of possible underlying mechanisms that may lead to “primary” RV dysfunction. For example, excess adipose accumulation has been associated with an expansion in both central and total blood volume, possibly through increased RAAS activation and aldosterone production, thereby also increasing preload which may lead to RV remodeling [26, 27]. In experimental mouse models, excess adiposity has also been shown to increase cardiovascular inflammation and cardiac aging [28, 29]. This effect appears to be modulated through macrophage infiltration, cardiac lipotoxicity from triglyceride accumulation in cardiac myocytes, and increased deposition of extracellular matrix proteins [28-31]. Another possibility is that RV dysfunction may be particularly significant in individuals with obesity as a “secondary” process from left-sided heart failure and pulmonary hypertension. For example, a study by Gorter et al examined 75 individuals with HFpEF and increased epicardial adipose tissue, finding that increased epicardial adipose tissue was associated with a higher BMI as well as higher right-sided filling pressures independent of pulmonary vascular resistance [32]. However, the stratified analyses we performed suggest an association between obesity and RV dysfunction that is present irrespective of pre-existing heart failure or pulmonary hypertension.

Our study has several limitations worth noting. First, this is a hospital-based study of ambulatory and hospitalized patients undergoing right heart catheterization for clinical indications, and both selection bias and confounding by indication may limit generalizability to other samples. Additionally, causal inferences could not be drawn due to the observational nature of the study design and the possibility of residual confounding. Another limitation is the use of code-based diagnoses of HF endpoints which introduce the possibility of misclassification. The potential limitations of right heart catheterization must also be considered as well, particularly in patients with obesity.

One study found that obesity confounds estimations of CO and PVR when calculated by the indirect Fick method due to a lower mixed venous oxygen saturation, potentially resulting in inappropriate hemodynamic classification [33]. To avoid this potential confounder, we utilized thermodilution rather than Fick where applicable (eg, with RVSWI).

In sum, individuals with obesity had worse hemodynamic indices of RV function, including lower PAPi and higher RAP:PCWP ratio across a broad hospital-based sample. While RV dysfunction was associated with worse clinical outcomes including mortality and HF hospitalization, this association was especially pronounced among individuals with higher BMI. Our findings highlight the important role of obesity in RV dysfunction and that future studies are needed to further understand the mechanism by which obesity leads to RV dysfunction and adverse outcomes. Further, with rapidly evolving anti-obesity therapies, potential clinical implications with respect to disease prevention are of high interest.

## Data Availability

All data produced in the present work are contained in the manuscript

## Acknowledgments

None

---

**Supplemental table 1.** Cross-sectional association with BMI and RV function stratified by prevalent HF

| Predictors | No prevalent HF (n=5629) |  |  | Prevalent HF (n=2656) |  |  |
| --- | --- | --- | --- | --- | --- | --- |
| | $\beta$ estimate | Standard error | p value | $\beta$ estimate | Standard error | p value |
| ***PAPi*** |  |  |  |  |  |  |
| BMI | -0.27 | 0.01 | <0.001 | -0.24 | 0.02 | <0.001 |
| ***RA:PCWP ratio*** |  |  |  |  |  |  |
| BMI | 0.26 | 0.01 | <0.001 | 0.25 | 0.02 | <0.001 |
| ***RVSWI*** |  |  |  |  |  |  |
| BMI | 0.03 | 0.02 | 0.03 | -0.003 | 0.02 | 0.89 |

**Supplemental table 2.** Cross-sectional association with BMI and RV function stratified by prevalent PH

| Predictors | No Prevalent PH (n=3077) |  |  | Prevalent PH (n=5208) |  |  |
| --- | --- | --- | --- | --- | --- | --- |
| | $\beta$ estimate | Standard error | p value | $\beta$ estimate | Standard error | p value |
| ***PAPi*** |  |  |  |  |  |  |
| BMI | -0.25 | 0.02 | <0.001 | -0.23 | 0.01 | <0.001 |
| ***RA:PCWP ratio*** |  |  |  |  |  |  |
| BMI | 0.28 | 0.02 | <0.001 | 0.24 | 0.01 | <0.001 |
| ***RVSWI*** |  |  |  |  |  |  |
| BMI | -0.08 | 0.02 | <0.001 | -0.04 | 0.01 | 0.004 |

**Supplement Figure 1.**
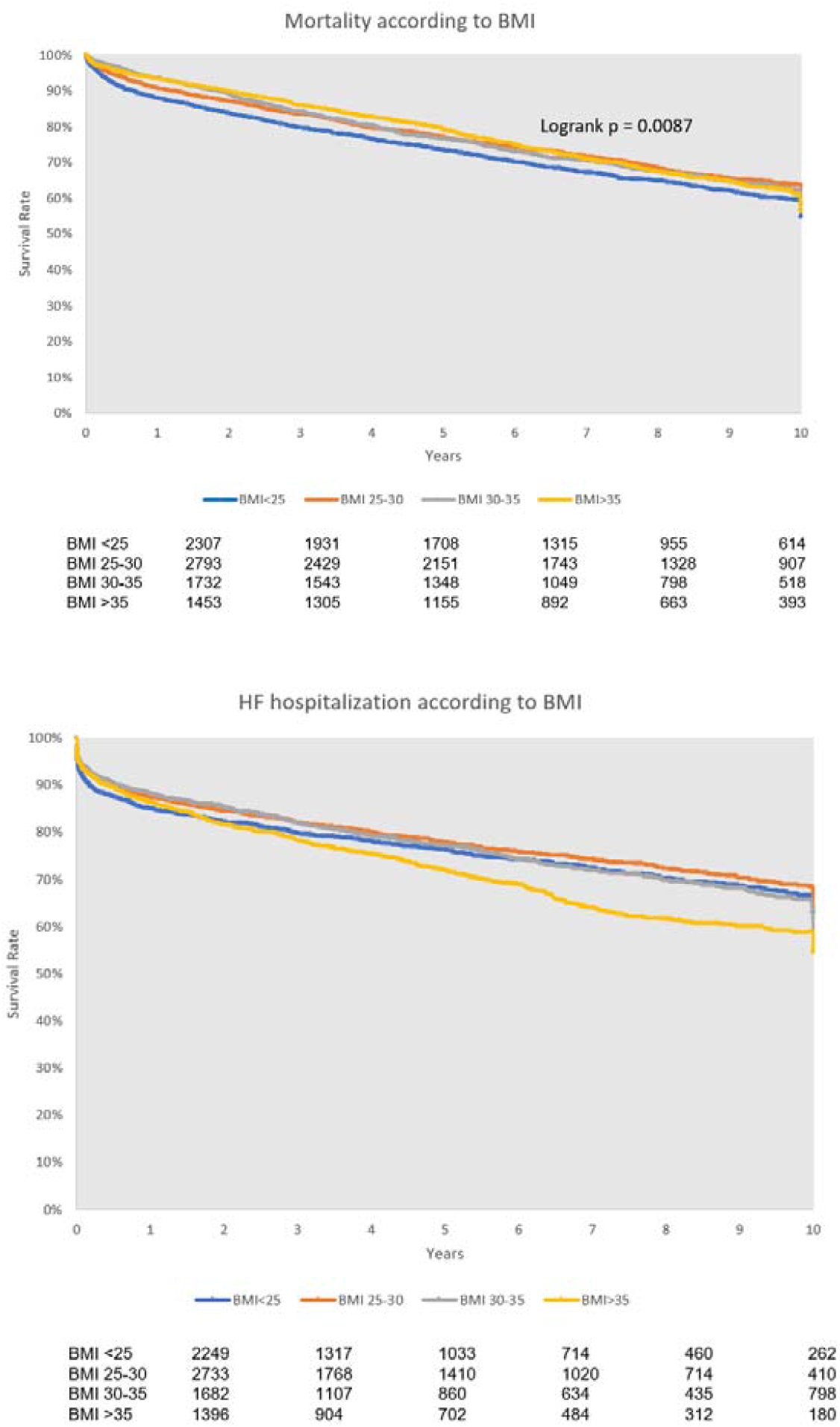
Kaplan-Meier survival curves across obesity classes

